# Cardiovascular disease and type 2 diabetes in older adults: a combined protocol for an individual participant data analysis for risk prediction and a network meta-analysis of novel anti-diabetic drugs

**DOI:** 10.1101/2023.03.13.23287105

**Authors:** Valerie Aponte Ribero, Heba Alwan, Orestis Efthimiou, Nazanin Abolhassani, Douglas C Bauer, Séverine Henrard, Antoine Christiaens, Gérard Waeber, Nicolas Rodondi, Baris Gencer, Cinzia Del Giovane

**Author notes:** Corresponding author **Correspondence to:** Valerie Aponte Ribero, Institute of Primary Health Care (BIHAM), University of Bern, Mittelstrasse 43, 3012 Bern, Switzerland. Joint first authors (Valerie Aponte Ribero, Heba Alwan).

## Abstract

**Introduction:** Older and multimorbid adults with type 2 diabetes (T2D) are at high risk of cardiovascular disease (CVD) and chronic kidney disease (CKD). Estimating risk and preventing CVD is a challenge in this population notably because it is underrepresented in clinical trials. Our study aims to (1) assess if T2D and haemoglobin A1c (HbA1c) are associated with the risk of CVD events and mortality in older adults, (2) develop a risk score for CVD events and mortality for older adults with T2D, (3) evaluate the comparative efficacy and safety of novel antidiabetics.

**Methods and analysis:** For Aim 1, we will analyse individual participant data on individuals aged ≥65 years from five cohort studies: the Optimising Therapy to Prevent Avoidable Hospital Admissions in Multimorbid Older People study; the Cohorte Lausannoise study; the Health, Aging and Body Composition study; the Health and Retirement Study; and the Survey of Health, Ageing and Retirement in Europe. We will fit flexible parametric survival models (FPSM) to assess the association of T2D and HbA1c with CVD events and mortality. For Aim 2, we will use data on individuals aged ≥65 years with T2D from the same cohorts to develop risk prediction models for CVD events and mortality using FPSM. We will assess model performance, perform internal-external cross validation, and derive a point-based risk score. For Aim 3, we will systematically search randomized controlled trials of novel antidiabetics. Network meta-analysis will be used to determine comparative efficacy in terms of CVD, CKD, and retinopathy outcomes, and safety of these drugs. Confidence in results will be judged using the CINeMA tool.

**Ethics and dissemination:** Aims 1 and 2 were approved by the local ethics committee (Kantonale Ethikkommission Bern); no approval is required for Aim 3. Results will be published in peer-reviewed journals and presented in scientific conferences.

**STRENGTHS AND LIMITATIONS:**

- We will analyse individual participant data from multiple cohort studies of older adults who are often not well represented in large clinical trials.
- By using flexible survival parametric models, we will be able to capture the potentially complex shapes of the baseline hazard functions of cardiovascular disease (CVD) and mortality.
- Our network meta-analysis will include recently published randomised controlled trials on novel anti-diabetic drugs that have not been included in previous network meta-analysis and results will be stratified by age and baseline HbA1c
- Although we plan to use several international cohorts, the external validity of our findings and particularly of our prediction model will need to be assessed in independent studies
- Our study will help guide CVD risk estimation and prevention among older adults with type 2 diabetes

## INTRODUCTION

Type 2 diabetes (T2D) is highly prevalent, affecting one in ten adults aged 20-79 years worldwide, and the prevalence rises to almost 25% in individuals aged 75-79 years [1, 2]. Cardiovascular (CVD) and chronic kidney diseases (CKD) are life-threatening complications of T2D. A large proportion of older individuals aged ≥65 years with T2D are multimorbid, where multimorbidity is defined as the presence of two or more chronic medical conditions [3]. Prevention of CVD and CKD is therefore critical in this population [4, 5].

In the following, we address three major open issues in the preventive care of adults with T2D: (1) the association of T2D and hemoglobin A1c (HbA1c) with the risk of CVD events and mortality in older adults; (2) the accurate prediction of CVD events and mortality risk in older adults with T2D; (3) the comparative efficacy and safety of novel antidiabetics to prevent CVD and CKD complications.

### Are T2D and HbA1c associated with the incidence of CVD events and mortality in older adults, and is T2D a coronary risk equivalent in this population?

It is uncertain whether T2D is independently predictive of incidence of CVD events and mortality in older and multimorbid adults, as studies have observed a decreasing association with older age [6-9]. A recent systematic review of studies evaluating CVD risk factors in people aged ≥60 years found that in two thirds of the studies, T2D was identified as a predictor of incident CVD [6]. Among studies with a mean age ≥75 years, however, only around one third retained T2D as a predictor for CVD in the final models.

It is also disputed whether T2D is a coronary heart disease (CHD) risk equivalent among older adults. Two large contemporary studies found a lower risk of developing CHD in diabetics without prior CHD compared to non-diabetics with previous CHD [10, 11]. Yet, two studies that were conducted exclusively in older adults reported a similar risk of CVD across these two groups, supporting the status of diabetes as a coronary risk equivalent in this population [12, 13].

Intensive glycemic control and diabetes overtreatment can result in harms such as increased risks of severe hypoglycemia and mortality, which may outweigh clinical benefits in older populations [14]. Therefore, recent guidelines recommend less stringent targets of HbA1c, and different targets according to individual’s health status, for older or multimorbid adults [15-18]. However, recommendations on optimal HbA1c targets are based on low-level evidence. Prospective observational studies that assessed the association of Hb1Ac with the risk of CVD and/or mortality among older adults with and without T2D reported conflicting results and were mostly limited to mortality outcomes [19-24].

Further assessment of these associations in a large sample of older and multimorbid adults is needed. A single cohort might have limited power to detect such an association. Analysing multiple cohorts increases power, precision, and might give insight into the heterogeneity of the association across different populations and settings. Analysis of individual participant data (IPD) allows for harmonization of analyses across studies and use of additional information that would not be possible with aggregate data, and is therefore the most powerful method for summarizing evidence from multiple cohorts.

### Can CVD events and mortality be accurately predicted in older adults with T2D?

Guidelines for the management of T2D recommend using risk scores to identify adults who are at high risk of CVD events [25]. However, most CVD risk-estimation tools developed for adults with T2D have not focused on older people [26], and recent external validation studies found that existing scores had poor predictive performance in older age groups [27, 28]. A risk score that can accurately predict the risk of CVD events and mortality in older and multimorbid adults is therefore needed. It is particularly relevant to identify, among older adults with T2D, those with a higher risk of CVD events, as they might benefit from medical treatment.

### Which novel anti-diabetic drug has the best benefit-risk profile for prevention of CVD and CKD, and should the medical management differ by age or glycemic control?

Novel anti-diabetic drugs have emerged, including sodium-glucose co-transporter-2 (SGLT-2) inhibitors, glucagon-like peptide 1 (GLP-1) receptor agonists (RA), and dipeptidyl peptidase-4 (DPP-4) inhibitors [29]. These drugs have been shown to have cardiovascular and renal benefits and have low risk of hypoglycemia [29]. However, recommendations differ according to the type of novel anti-diabetic drug. For example, the European Society of Cardiology (ESC) and the European Association for the Study of Diabetes (EASD) recommend treating patients with prevalent CVD or at high/very high CVD risk with either empagliflozin or liraglutide to reduce CVD events, whereas it is recommended to treat patients with SGLT2 inhibitors rather than GLP-1 RA to reduce the risk of hospitalization due to heart failure [30, 31]. Moreover, the therapeutic effect of novel antidiabetic drugs may vary between drugs, between younger and older adults [32], and according to baseline HbA1c levels [33]. A network meta-analysis (NMA) can be used to make such comparisons, utilizing all available data [34]. Only a few NMAs have compared the classes of novel anti-diabetic drugs with each other in terms of preventing CVD, CKD or mortality [35-40]. None of the published NMAs have provided results according to baseline levels of HbA1c and only one reported analysis stratified in younger and older adults separately [39].

It is therefore timely to conduct an up-to-date, comprehensive, and high-quality NMA to assess, overall and according to age and baseline HbA1c levels, the benefit and safety profile of novel anti-diabetic drugs in adults with T2D.

Our overarching goal is to improve CVD risk prediction among multimorbid older adults with T2D and to compare the benefits and harms of novel anti-diabetic drugs. We endeavour to accomplish this goal with three specific aims:

1. To assess the association of i) T2D and ii) different HbA1c levels with the risk of CVD events and mortality, as well as to assess if iii) T2D is a CHD equivalent in older adults, including people with multimorbidity.
2. To develop a novel score for predicting the 5- and 10-year risks of CVD events and mortality in older adults with T2D, including people with multimorbidity.
3. To evaluate the comparative efficacy and safety of novel antidiabetic drugs in individuals with T2D in the overall population, in younger and older patients, and for different baseline HbA1c levels.

Methods are described separately for each aim in the following section.

## METHODS AND ANALYSIS

### Patient and public involvement

Patients or the public were not involved in the design, or conduct, or reporting, or dissemination plans of our research.

### Aim 1: Assessing the association of T2D and HbA1c with the risk of CVD events and mortality, and evaluating if T2D is a coronary risk equivalent, in older adults

All results will be reported in accordance with the Strengthening the Reporting of Observational Studies in Epidemiology (STROBE) guideline for cohort studies [41].

#### Study design and participants

For Aim 1 of this study, we will use IPD on participants aged ≥65 years from five prospective cohort studies (Table 1): the Optimising Therapy to Prevent Avoidable Hospital Admissions in Multimorbid Older People (OPERAM) study [42, 43]; the Cohorte Lausannoise (CoLaus) study [44]; the Health, Aging, and Body Composition (Health ABC) study [45]; the Health and Retirement Study (HRS) [46]; and the Survey of Health, Ageing and Retirement in Europe (SHARE) [47, 48]. Study descriptions are available in the Supplementary Appendix S1.

**Table 1.**
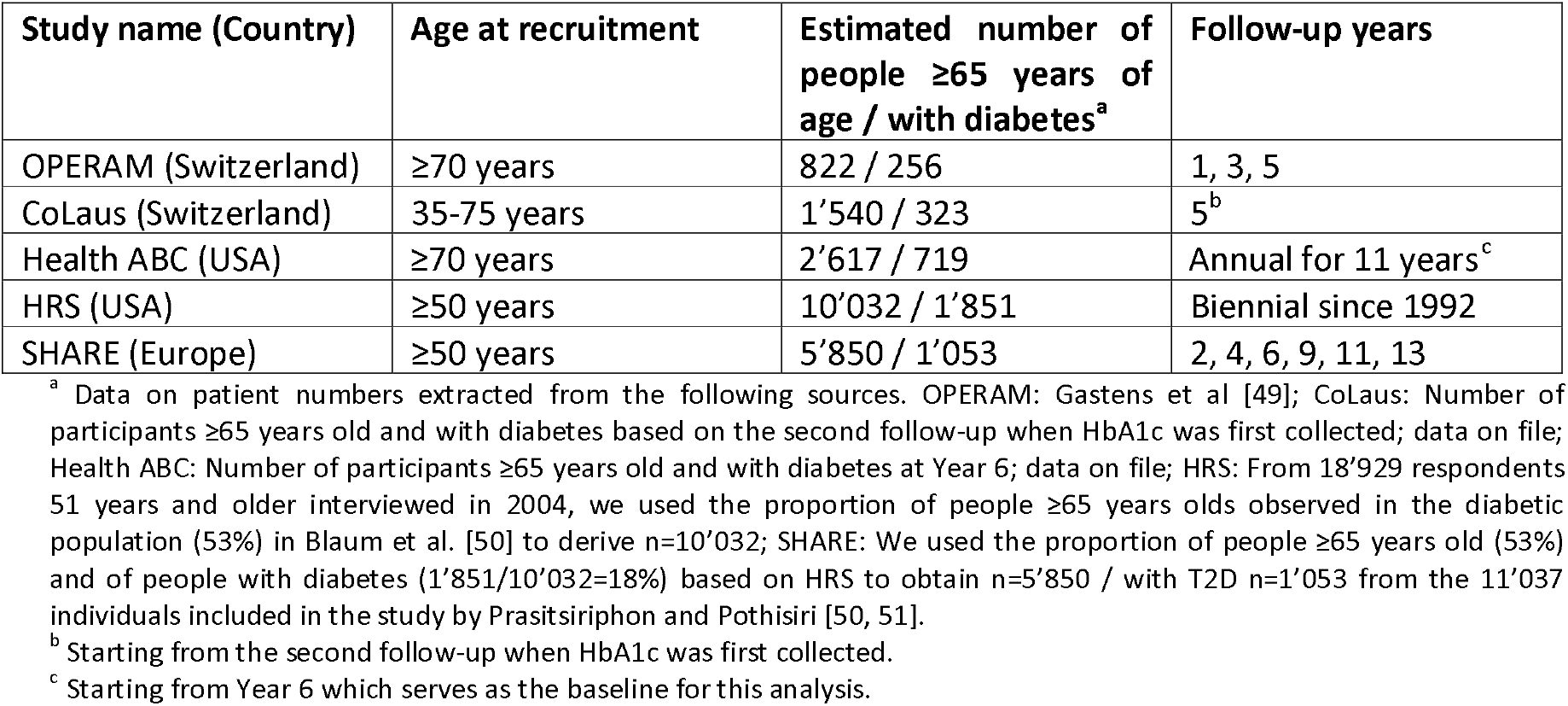
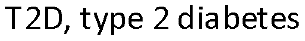
List of cohort studies to be included in the analysis

| Study name (Country) | Age at recruitment | Estimated number of people $\geq 65$ years of age / with diabetes<sup>a</sup> | Follow-up years |
| --- | --- | --- | --- |
| OPERAM (Switzerland) | $\geq 70$ years | 822 / 256 | 1, 3, 5 |
| CoLaus (Switzerland) | 35-75 years | 1'540 / 323 | 5<sup>b</sup> |
| Health ABC (USA) | $\geq 70$ years | 2'617 / 719 | Annual for 11 years<sup>c</sup> |
| HRS (USA) | $\geq 50$ years | 10'032 / 1'851 | Biennial since 1992 |
| SHARE (Europe) | $\geq 50$ years | 5'850 / 1'053 | 2, 4, 6, 9, 11, 13 |

<sup>a</sup> Data on patient numbers extracted from the following sources. OPERAM: Gastens et al [49]; CoLaus: Number of participants $\geq 65$ years old and with diabetes based on the second follow-up when HbA1c was first collected; data on file; Health ABC: Number of participants $\geq 65$ years old and with diabetes at Year 6; data on file; HRS: From 18'929 respondents 51 years and older interviewed in 2004, we used the proportion of people $\geq 65$ years olds observed in the diabetic population (53%) in Blaum et al. [50] to derive $n=10'032$ ; SHARE: We used the proportion of people $\geq 65$ years old (53%) and of people with diabetes ( $1'851/10'032=18\%$ ) based on HRS to obtain $n=5'850$ / with T2D $n=1'053$ from the 11'037 individuals included in the study by Prasitsiripon and Pothisiri [50, 51].
<sup>b</sup> Starting from the second follow-up when HbA1c was first collected.
<sup>c</sup> Starting from Year 6 which serves as the baseline for this analysis.
T2D, type 2 diabetes

Data from all participants with and without T2D aged ≥65 years at baseline will be included for this analysis. We expect to include data from at least 20’861 participants (Table 1). Final data on the number of patients included in the analyses will be determined once data collection has been completed.

#### Exposures

The exposures of interest are (i) presence of T2D at baseline, (ii) HbA1c levels at baseline, and (iii) no T2D and prior CHD versus T2D and no prior CHD at baseline. Definitions are available in the Supplementary Appendix S2.

#### Outcomes

The primary outcome is a composite of incidence of CVD event or all-cause death. We decided to include all-cause death in the primary outcome rather than CVD-related death only, as mortality is high in older individuals, with various causes leading to death, and focusing on CVD-related death alone might exaggerate a potentially unimportant safety signal. Secondary outcomes will be the individual components of the composite: incidence of a fatal or non-fatal CVD event, and all-cause death.

The outcome of CVD event will be defined according to the cohort study included. We will use data from the entire available follow-up of participants. Outcomes will be censored at the time of any CVD event, death, or the last follow-up assessment (whichever comes first). If multiple CVD events have been observed, only the first will be considered. Adjudicated outcomes will be used in the analyses whenever possible. Unadjudicated outcomes will be included when no adjudication was performed in the study. A summary of the adjudication procedures across the studies is provided in the Supplementary Appendix S3.

#### Statistical analysis

We will perform an IPD analysis on the five cohort studies using a two-step approach, in which models will first be fit to each study separately and results subsequently meta-analysed [52, 53]. We will use a flexible survival parametric model to analyse the association between each outcome and i) T2D (as a dichotomous variable), and ii) HbA1c levels (as a continuous variable), and estimate the respective hazard ratios and 95% confidence intervals (CIs) [54]. For the secondary outcome on fatal and non-fatal CVD event, we will use a competing-risk model with non-CVD-related deaths as a competing event and estimate sub-hazard ratios [55]. A potential non-linear relationship for continuous variables such as HbA1c will be accounted for by including splines in the model. In order to determine if T2D is a coronary risk equivalent among multimorbid older adults, we will compare the risk of CVD events and death of non-diabetic adults who had a previous CHD event to that of diabetic adults with no prior CHD [12].

We will perform an analysis adjusted for baseline age and sex. We will further adjust our models for the following covariates: body mass index, prior CVD, smoking status, alcohol consumption, systolic blood pressure, hypertension treatment, total cholesterol, high-density lipoprotein cholesterol, and treatment for high cholesterol.

Subgroup analyses will be conducted to detect effect modification or significant interaction terms that need to be included in the model. Subgroups will be age (≥ 75 years vs < 75 years), sex (women vs men) and prior CVD (yes vs no). For the assessment of the predictive value of HbA1c, we will also perform stratified analyses by presence of T2D at baseline. In individuals with baseline T2D, we will conduct a subgroup analysis by treatment with hypoglycaemic medications (including insulins, glinides and sulfonylureas; yes vs no) if sufficient data is available, and a sensitivity analysis in which HbA1c will be categorized as <7.5%, ≥7.5% to <8.4% and ≥ 8.5% [15].

We will use multiple imputation methods to impute missing data for the analyses [56, 57].

#### Power estimation

To assess if our sample size is sufficient, we calculated the power to detect an increased risk of the primary outcome (CVD event and overall mortality) in older people with T2D. We varied the risks at 5-year follow-up in older people without T2D between 12 to 25% with a minimum relative risk of 1.2 for individuals with versus without T2D, based on 1) the mortality risk at 1-year follow-up in OPERAM that was (18%) [43], and 2) the mortality risk at 5-years follow-up from the Cardiovascular Health Study (12%) [58], respectively. Given an expected sample size of 20’861 and 4’202 adults without and with T2D, respectively, power is at least 99% (alpha = 0.05; two-sided test).

### Aim 2: Development of risk-prediction models for CVD events and mortality in older adults

Results from all analyses will be reported according to the Transparent Reporting of a multivariable prediction model for Individual Prognosis Or Diagnosis statement (TRIPOD) guidelines [59].

#### Study design and participants

For the development of the CVD events and mortality risk prediction model, we will use IPD on the subgroup of participants aged ≥65 years with T2D at baseline from the same sources as in Aim 1 (Table 1). The estimated sample size available for this analysis is 4’202 individuals with T2D.

#### Outcomes

The primary and secondary outcomes will be the same as for Aim 1. We will estimate the 5- and 10-year risks of these outcomes.

#### Statistical analysis

For the development of the risk-prediction models, we will fit flexible survival parametric models to the IPD from the cohort studies using a one-step approach [54, 60]. We will assess the heterogeneity of baseline risk and predictor effects as recommended by Debray et al. [60]. If baseline risks are not considered to be very different across the cohorts, we will derive a single model with a random intercept using IPD from all cohorts [61]. Otherwise, we will estimate study-specific intercepts and give guidance on choosing the most appropriate intercept for a population [62]. We will use splines to model potential non-linear relationships between continuous variables and the outcome. For the secondary outcome of fatal and non-fatal CVD event, we will use a competing-risk model [55, 63]. We will use multiple imputation to impute missing data for the analyses [56, 57].

#### Predictor selection

Using predictors that have a causal relationship with the outcome may improve transportability of clinical prediction models [64]. Therefore, we will map the causal relationship between potential candidate predictors and CVD events and mortality using directed acyclic graphs (DAGs). Potential candidate predictors were identified from previously reported CVD risk scores for individuals with T2D [28], listed in Table 2, and will be collected from all five data cohorts if available. Final predictors to be included in the model will be based on the DAGs, clinical guidance regarding practical usability of the model, and availability across the cohorts.

**Table 2.**
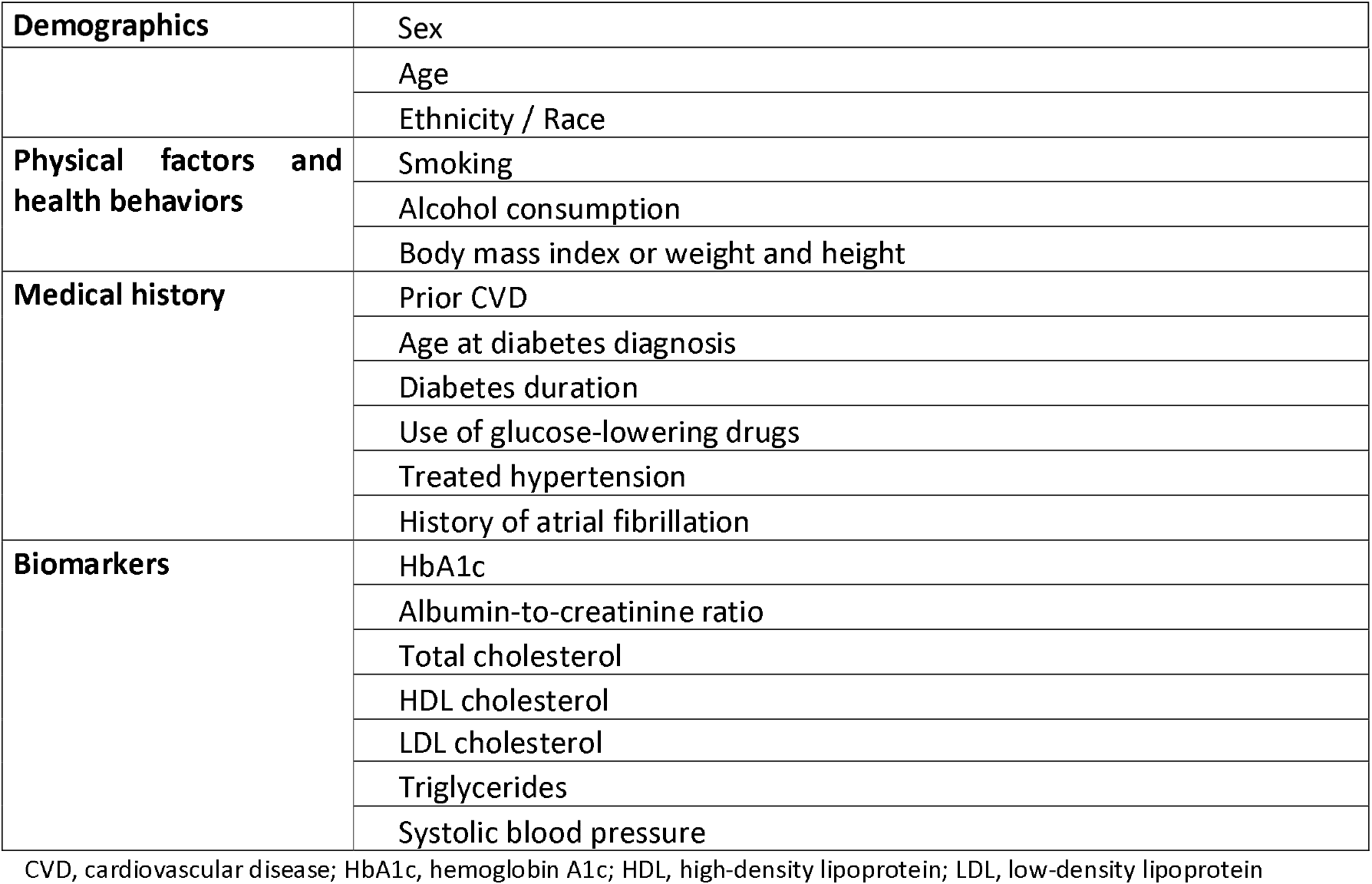
Collection of baseline characteristics from cohort studies

|  |  |
| --- | --- |
| Demographics | Sex |
|  | Age |
|  | Ethnicity / Race |
| **Physical factors and health behaviors** | Smoking |
|  | Alcohol consumption |
|  | Body mass index or weight and height |
| **Medical history** | Prior CVD |
|  | Age at diabetes diagnosis |
|  | Diabetes duration |
|  | Use of glucose-lowering drugs |
|  | Treated hypertension |
|  | History of atrial fibrillation |
| **Biomarkers** | HbA1c |
|  | Albumin-to-creatinine ratio |
|  | Total cholesterol |
|  | HDL cholesterol |
|  | LDL cholesterol |
|  | Triglycerides |
|  | Systolic blood pressure |

CVD, cardiovascular disease; HbA1c, hemoglobin A1c; HDL, high-density lipoprotein; LDL, low-density lipoprotein

#### Assessing model performance and model validation

The model’s performance will be assessed using measures for discrimination, calibration, and overall performance. For discrimination, we will calculate Harrell’s concordance (c) statistic for time-to-event data [65]. Calibration will be assessed using calibration plots, the Greenwood-D’Agostino-Nam test of calibration and expected-to-observed ratio [66, 67]. Overall performance of the model will be evaluated using Nagelkerke’s R^2^ [66].

We will validate the model internally using bootstrapping to calculate optimism-corrected c-statistics [68]. We will also follow an internal-external cross-validation method [60]. This method allows us to examine differences in the performance of the model across studies and assess the generalizability and applicability of the developed model. Based on these validation exercises, we may make adjustments to the risk-prediction model (e.g, by excluding strongly heterogeneous predictors, or by including interaction effects) [60].

#### Calculation of a novel risk score and implementation of an online risk prediction tool

We will develop an online risk-prediction tool of the final model using the Shiny R package [69]. If a model including only linear predictors provides good predictive performance, we will also create a point-based risk scoring system to facilitate clinical use of the risk-prediction model. We will assign integer points to each predictor and predictor level of the final model, according to the system of the Framingham risk score [70].

#### Sample size estimation

We calculated the required sample size for the risk-prediction model [71]. Assuming an outcome prevalence of 15%, an R^2^ of 15% in the primary outcome, and 19 predictors (assuming no splines), the minimum sample size required for a new model development is 1’043 participants (corresponding to 8 events per predictor parameters [EPP]). For an R^2^ of 10%, a sample size of 1’614 would be required. For an outcome prevalence of 25% (with an R^2^ of 15%), the sample size is the same with a higher EPP of 14. Therefore, our sample size of 4’202 is deemed adequate.

### Aim 3: Systematic review and NMA to evaluate the comparative efficacy and safety of novel antidiabetic drugs in individuals with T2D

This systematic review and NMA is registered in the International Prospective Register of Systematic Reviews (PROSPERO; CRD 42022310243). We will adhere to the Preferred Reporting Items for Systematic reviews and Meta-Analyses (PRISMA) extension statement for reporting of systematic reviews incorporating network meta-analyses of healthcare interventions [72].

#### Eligibility criteria

Studies that meet the following criteria will be included: (1) randomized controlled trials that included adults 18 years or older with T2D; (2) studies that included at least one of the following novel anti-diabetic drugs: SGLT-2 inhibitors, GLP1-RA, and DPP-4 inhibitors; (3) studies that included a control group of either placebo, no drug, another novel anti-diabetic drug, or older anti-diabetic drugs (metformin, insulin secretagogues, alpha-glucosidase inhibitors, and thiazolidinediones). Studies that only included older anti-diabetic drugs and compared them with each other or placebo will be excluded.

#### Types of outcome measures

##### Primary outcomes

The primary outcomes will be (1) incidence of major adverse cardiovascular events (MACE), defined as the composite of cardiovascular mortality, nonfatal myocardial infarction (MI), and nonfatal stroke; (2) renal composite outcome as defined in each trial, such as a composite of adjudication-confirmed end-stage renal disease (ESRD), death due to renal failure, new onset macro-albuminuria, or a sustained decrease of at least 40% in estimated glomerular filtration rate from baseline to less than 60 ml per minute per 1.73 m^2^ of body-surface area [31, 73]; and (3) diabetic retinopathy as defined by each trial, including vitreous haemorrhage, onset of diabetes-related blindness, or the need for retinal photocoagulation [74].

##### Secondary outcomes

Secondary outcomes will include CV mortality; heart failure; myocardial infarction (fatal and non-fatal); coronary and/or peripheral revascularization; all strokes (fatal and non-fatal); all-cause mortality; and HbA1c level measured at follow-up. The following safety outcomes will be assessed: proportion of participants with at least one serious adverse event (e.g., severe hypoglycemia, lower limb amputation, bone fracture, and diabetic ketoacidosis); proportion of participants with a specific serious adverse event; and proportion of participants who withdrew due to adverse events, such as hypoglycemia.

#### Search Strategy

##### Information sources

The following databases will be searched for eligible studies: MEDLINE, Embase, Cochrane Library, and clinical trial registries (http://clinicaltrials.gov/ and the World Health Organization). We will hand-search the reference lists of all articles, texts, and other reviews on the topic we retrieved, and contact authors and researchers active in the field for more data. We will not apply language and time restrictions to our search.

##### Identification and selection of studies

Two researchers will independently select studies, extract and collect data in a two-step process. First, we will screen the titles and abstracts. Second, we will read the full texts of all potentially relevant studies and determine the final list of studies to include. When discrepancies arise that cannot be resolved by consensus between the two researchers, a third senior author will be consulted.

#### Data extraction

Two reviewers will extract data into pre-specified data extraction forms [75]. For each study, we will extract information on study characteristics (e.g., setting, study design, sample size, follow-up), participant characteristics (e.g., age, sex, duration of diabetes, BMI, presence of comorbidities, previous CVDs, baseline HbA1c levels), interventions and controls (e.g., dose, frequency of intervention) and outcomes. For binary outcomes, we will extract the number of patients with the event, the relative risk, odds ratio and hazard ratio and their CIs. We may consider combining hazard ratio and relative risk. For continuous outcomes, when follow-up data are not reported and only change from baseline is available, we will use the latter [76]. We will use published standard deviation (SD), where available. If SD are not available from the publication, SD will be calculated from p values, t-values, CIs or standard errors [77].

#### Risk of bias assessment

We will use the Cochrane Collaboration ‘risk of bias’ tool to assess risk of bias (RoB) for each included study [78, 79]. Bias will be evaluated in the following five domains: (1) sequence generation (2) allocation concealment, (3) blinding of participants, personnel, and outcome assessors, (4) incomplete outcome data, and (5) selective outcome reporting. Studies will be classified as having a high, low, or unclear risk of bias overall and for each of the five domains. Two reviewers will independently assess the risk of bias in selected studies. Disagreements will be resolved by discussion and, if needed, by consulting a third senior author.

#### Assessing clinical and methodological heterogeneity within and across comparisons of drugs

In each pairwise comparison, patient characteristics, drugs and outcome definitions of included studies should be similar [78]. We will produce descriptive statistics for studies and assess their similarity in each comparison. If the assumption of transitivity can be defended, [34] we will compare the distribution of the potential effect modifiers across the different pairwise comparisons [80, 81]. We will assess transitivity for the following possible effect modifiers: dose, frequency or duration of drug, diabetes duration at baseline, sex, high vs low cardiovascular risk trials, baseline levels of HbA1c, and risk of bias. If we find evidence of important differences across comparisons, we will explore the effects of potential effect modifiers with network meta-regression or subgroup analysis. We assume that all treatments are jointly randomizable.

#### Data analysis

For each outcome, we will conduct pairwise random-effects meta-analyses. Pooled relative effects will be shown along with their 95% CIs. If transitivity is deemed plausible, we will perform a random effects NMA including all studies. For drug ranking, we will use the P-score to provide a hierarchy for each outcome separately and a revised version of the P-score that accounts for multiple outcomes and of the clinical importance value. This will allow us to assess how much harm can be tolerated for a certain benefit [82]. We will estimate the variance of random effects for each pairwise comparison in standard pairwise meta-analyses and assess the magnitude of heterogeneity by visually inspecting the forest plots, by calculating prediction intervals and calculating the I^2^ statistic [83]. We will assess the magnitude of heterogeneity by comparing the estimated value with empirical distributions, and by examining prediction intervals [84, 85]. Statistical disagreement between direct and indirect effect sizes will be evaluated both using local (node-splitting) and global approaches (design-by-treatment test) [83].

We will assess the existence of small study effects and publication bias with a contour-enhanced funnel plot for each pairwise comparison with more than 10 studies, and by running Egger’s test. For rare outcomes, we will use a NMA model for rare events [86].

We will use STATA version 16 software and R for our analysis (http://methods.cochrane.org/cmi/network-meta-analysis-toolkit) [87, 88]. NMA results will be presented in league tables and forest plots [87]. We will present trade-offs between benefits and harms for each treatment in a two-dimensional plot. We will judge the confidence in the evidence derived from NMA with the CINeMA (http://cinema.ispm.ch/) tool [89, 90]. We will stratify analysis according to low versus high HbA1c levels, by age (more or less than 65 years), and cardiovascular risk trials (high vs low). We will perform sensitivity analysis by excluding a) trials with less than 12 months of follow-up and b) studies with risk of bias.

## Data Availability

All data produced in the present work are contained in the manuscript

## ETHICS AND DISSEMINATION

Aims 1 and 2 of this study were approved by the local ethics committee (Kantonale Ethikkommission Bern). No ethics approval was required for Aim 3. The results of this study will be published within multiple articles in peer-reviewed journals and presented in meetings.

### Contributorship statement

CDG, NR, GW and BG conceived the original idea, study concept and design. CDG, NR, BG, VAR, HA and OE contributed to the study design. VAR and HA drafted the manuscript and contributed equally to this manuscript. All authors critically revised the manuscript for important intellectual content.

### Funding

This work is supported by the Swiss National Science Foundation grant number (325130_204361 / 1 to Dr. Baris Gencer and Prof. Gérard Waeber).

This work is part of the project “OPERAM: OPtimising thERapy to prevent Avoidable hospital admissions in the Multimorbid elderly” supported by the European Union’s Horizon 2020 research and innovation programme under the grant agreement No 6342388, and by the Swiss State Secretariat for Education, Research and Innovation (SERI) under contract number 15.0137. The opinions expressed and arguments employed herein are those of the authors and do not necessarily reflect the official views of the EC and the Swiss government.

This work uses data from the Health, Aging and Body Composition Study supported by National Institute on Aging (NIA) Contracts N01-AG-6-2101; N01-AG-6-2103; N01-AG-6-2106; NIA grant R01-AG028050, and NINR grant R01-NR012459. This study was funded in part by the Intramural Research Program of the NIH, National Institute on Aging.

This work uses data from the CoLaus|PsyCoLaus Study. The CoLaus study was supported by research grants from GlaxoSmithKline, the Faculty of Biology and Medicine of Lausanne, Switzerland and the Swiss National Science Foundation (grants no: 3200B0–105993, 3200B0-118308, 33CSCO-122661, 33CS30-139468, 33CS30-148401 and 33CS30_177535).

This work uses data from SHARE Waves 1, 2, 3, 4, 5, 6, 7, and 8 (DOIs: 10.6103/SHARE.w1.800, 10.6103/SHARE.w2.800, 10.6103/SHARE.w3.800, 10.6103/SHARE.w4.800, 10.6103/SHARE.w5.800, 10.6103/SHARE.w6.800, 10.6103/SHARE.w7.800, 10.6103/SHARE.w8.800, 10.6103/SHARE.w8ca.800), see Börsch-Supan et al. (2013) for methodological details.

The SHARE data collection has been funded by the European Commission, DG RTD through FP5 (QLK6-CT-2001-00360), FP6 (SHARE-I3: RII-CT-2006-062193, COMPARE: CIT5-CT-2005-028857, SHARELIFE: CIT4-CT-2006-028812), FP7 (SHARE-PREP: GA N°211909, SHARE-LEAP: GA N°227822, SHARE M4: GA N°261982, DASISH: GA N°283646) and Horizon 2020 (SHARE-DEV3: GA N°676536, SHARE-COHESION: GA N°870628, SERISS: GA N°654221, SSHOC: GA N°823782, SHARE-COVID19: GA N°101015924) and by DG Employment, Social Affairs & Inclusion through VS 2015/0195, VS 2016/0135, VS 2018/0285, VS 2019/0332, and VS 2020/0313. Additional funding from the German Ministry of Education and Research, the Max Planck Society for the Advancement of Science, the U.S. National Institute on Aging (U01_AG09740-13S2, P01_AG005842, P01_AG08291, P30_AG12815, R21_AG025169, Y1-AG-4553-01, IAG_BSR06-11, OGHA_04-064, HHSN271201300071C, RAG052527A) and from various national funding sources is gratefully acknowledged (see www.share-project.org).

The HRS (Health and Retirement Study) is sponsored by the National Institute on Aging (grant number NIA U01AG009740) and is conducted by the University of Michigan.

## Acknowledgements

We thank Professor Arnaud Chiolero for his contribution to the development of the research protocol.

## Ethics approval

The study protocol was approved by the local ethics committee in Bern, Switzerland (Kantonale Ethikkommission Bern).

