## Supplementary appendix for "Cardiovascular disease and type 2 diabetes in older adults: a combined protocol for an individual participant data analysis for risk prediction and a network meta-analysis of novel anti-diabetic drugs"

### S1. Description of cohort studies included for Aims 1 and 2

OPERAM is a multi-center randomized controlled trial that was designed to evaluate whether a computer software - the Systematic Tool to Reduce Inappropriate Prescribing assistant (STRIPA) - can reduce drug-related hospital admissions (primary outcome) at 1-year follow-up in participants 70 years of age and older with multimorbidity (three or more chronic conditions) and polypharmacy (five or more regular medications) (1, 2). We will use data from the Bern site only for this study, for which follow-up has been extended to 5 years.

CoLaus is a single-center, population-based cohort study to investigate the epidemiology and genetic determinants of cardiovascular risk factors and metabolic syndrome starting in 2003 of patients aged 35 to 75 years living in Lausanne (3).

Health ABC is a prospective longitudinal study on risk factors for the decline of function and body composition with age in healthier older persons aged 70 years and older in the USA (4).

HRS is a longitudinal cohort study of more than 37,000 individuals over age 50 in 23,000 households in the USA (5). The survey has been fielded every 2 years since 1992. Since 2006, data collection has expanded to include biomarkers such as HbA1c.

SHARE is a cross-national panel study providing longitudinal micro data on health, socio-economic status, and social networks of about 140,000 individuals aged 50 or older in 27 European countries and Israel (6). Seven waves of data are currently available. Biomarker data has been collected from 24,000 individuals in Wave 6 (2014/15).

### S2. Definition of exposures in Aim 1

Type 2 diabetes will be defined as any of the following: 1) Self-reported diabetes; 2) Diabetes ascertained from medical records; 3) Diabetes medication use; 4) If blood test available: criteria for diabetes defined according to the American Diabetes Association (i.e. HbA1c >6.5%, fasting glucose ≥7 mmol/l or oral glucose tolerance test measurement ≥11 mmol/l) (7).

Coronary heart disease will be defined according to the information collected in the study and may include: 1) Self-reported heart disease; 2) Heart disease ascertained from medical records.

S3. Description of adjudication procedures across the cohort studies

In OPERAM, information on cardiovascular events and relative date for long follow-up will be collected by a nurse and identified according to the ICD-10 codes. Cardiovascular outcome will be adjudicated and centrally validated by an independent endpoint committee. Information on death and relative date will be collected through follow-up calls with the patients or GP contact.

In CoLaus, information on cardiovascular disease (CVD) and cause of death was collected from medical records or the Swiss National Death Registry and diagnosis of CVD was adjudicated by an adjudication committee according to international recommendations.

In Health ABC, all reported deaths were adjudicated through the end of data collection (30 September 2014), while adjudication for non-fatal events is only complete for events reported through 14 April 2012. Adjudication protocols are available online (https://healthabc.nia.nih.gov/disease-outcomes).

In HRS and SHARE, cardiovascular events and death were collected based on self- and proxy-report. No adjudication was conducted.
